# A Novel Dry-Stabilized Whole Blood Microsampling and Protein Extraction Method for Testing of SARS-CoV-2 Antibody Titres

**DOI:** 10.1101/2022.08.31.22279395

**Authors:** Patrick McCarthy, Joseph A Pathakamuri, Daniel Kuebler, Jocelyn Neves, Madison Krohn, Michael Rohall, Isaac Archibeque, Heidi Giese, Martina Werner, Ulrich Thomann

**Affiliations:** Covaris, LLC, Woburn, Massachusetts, USA; Department of Biology, Franciscan University, Steubenville, Ohio, USA

**Keywords:** COVID-19, SARS-CoV-2, anti-SARS-CoV-2 IgG, Adaptive-Focused Acoustics, ELISA, fingerstick blood, truCOLLECT whole blood, decentralised blood collection

## Abstract

The COVID-19 pandemic has revealed a crucial need for rapid, straightforward collection and testing of biological samples. Serological antibody assays can analyse patient blood samples to confirm immune response following mRNA vaccine administration or to verify past exposure to the SARS-CoV-2 virus. While blood tests provide vital information for clinical analysis and epidemiology, sample collection is not uncomplicated. This process requires a visit to the doctor’s office, a professionally trained phlebotomist to draw several millilitres of blood, processing to yield plasma or serum, and necessitates appropriate cold chain storage to preserve the specimen. The Covaris truCOLLECT Whole Blood Collection Kit allows for a lancet-based, decentralised capillary blood collection of exactly metered low volumes and eliminates the need for refrigerated transport and storage through the process of active desiccation. Anti-SARS CoV-2 spike and nucleocapsid protein antibody titres in plasma samples collected via venepuncture were compared to antibody titres in whole blood extracts obtained by treating desiccated whole blood samples stored in truCOLLECT sampling devices with Covaris Adaptive-focused Acoustics (AFA). Pearson correlation coefficients of 0.98, 95% CI [0.96, 0.99] for anti-SARS-CoV spike protein antibodies and 0.97, 95% CI [0.94, 0.99] for anti-SARS-CoV-2 nucleocapsid protein antibodies were observed. These data suggest that serology testing using desiccated whole blood samples collected and stored in truCOLLECT devices can be a convenient and cost-effective alternative to conventionally collected plasma.

## Introduction

The global Coronavirus Disease 2019 (COVID-19) pandemic has had a profound effect on industry, healthcare, and the individual due to its unprecedented impact on routine events. Exposure to the Severe Acute Respiratory Syndrome Coronavirus (SARS-CoV-2) generally results in symptoms 2 to 14 days after infection including fever, cough, shortness of breath, and loss of taste or smell [1]. Infection can be fatal or result in ongoing health problems that last for weeks or years such as chronic fatigue, heart palpitations, and difficulty thinking or concentrating [2]. Symptoms and severity of COVID-19 can be mitigated through the administration of either an mRNA or viral vector vaccine which confers resistance against SARS-CoV-2 by generating an immune response against the viral spike (S) glycoprotein [3]. Viral infection will generate an immune response against S protein as well but will additionally result in antibodies targeting other viral proteins such as the nucleocapsid (N) core [4]. This distinction enables blood testing to address a range of needs such as immunity conferred by vaccine or natural infection in both the clinical space as well as in serologic studies. As of July 2022, the World Health Organization (WHO) has reported an excess of 550 million confirmed cases of COVID-19, 6 million deaths, and 12 billion vaccine doses administered [5].

Contrary to PCR tests which are used to identify ongoing infection with SARS-CoV-2, serology tests are employed to provide information about vaccination efficiency, previous exposure, and severity of disease [6]. The Centers for Disease Control and Prevention (CDC) are utilizing these antibody tests as a means of surveillance to investigate how the virus is spreading throughout populations in the United States, and to learn more about the epidemiology of COVID-19 [7]. Serology testing has been used successfully to diagnose or determine immunity to other diseases including rabies [8], hepatitis [9], and West Nile Virus [10]. They are also used to measure antibody titres for the purpose of monitoring ongoing disease states [11]. Regardless of the assay, sample collection for serological tests requires a professionally trained phlebotomist to draw several millilitres of blood using highly invasive methods, rapid on-site processing to yield plasma or serum, and necessitates cold chain storage to preserve unstable biomarkers [12]. This process precludes self-collection and conventional transport/delivery by mail, results in sample waste due to collection of significantly more blood than needed, and requires additional processing steps such as plasma separation which together result in an unnecessary impediment to serological assays and overall cost burden.

To prepare for future surveillance of outbreaks and meet the rising interest in decentralised and convenient sample collection for serology testing, alternative methods of blood collection must be developed. Microsampling is an attractive strategy due to its reduction of blood collection volumes below 100 µL, however current techniques such as dried blood spot (DBS) cards and dried fingerstick blood might suffer from several challenges including difficult sample extraction and high variation between replicates due to unmetered specimen volumes [13, 14]. Microsampling of discrete volumes is crucial for accurate analysis and high performance of samples collected with a solid matrix [15]. Several commercial microsampling devices are available which rely on DBS-based or unstabilised liquid storage, and most are not designed for the extraction of large biomolecules [16]. Erstwhile attempts to iterate on current microsampling methods have resulted in poor correlations, indeterminant results, or the need for additional processing steps such as centrifugation or extended incubation periods [17–20].

The truCOLLECT Whole Blood Collection Kit was designed to address these challenges, by enabling decentralised whole blood collection, reducing collected blood volumes, eliminating the need for cold chain shipping and storage, and by circumventing the processing steps required to generate plasma or serum. Capillary blood is collected from a lanced finger via a K2-EDTA-coated capillary into a collection tube containing an immobilization reagent before undergoing active desiccation to preserve the sample for extended periods of time at ambient temperature (Figure 1). The immobilization matrix in the truCOLLECT sampling device was specifically designed to allow extraction of proteins as well as nucleic acids from dry-stabilized whole blood. After attaching a desiccation module, the immobilized blood sample can be shipped at ambient temperature while it is undergoing complete desiccation and hence stabilization. Extraction of biomolecules such as proteins and DNA from the desiccated blood sample is then performed on demand by adding an appropriate buffer and by subjecting the sample to Adaptive-Focused Acoustics (AFA), thereby actively hydrating and homogenizing the dried blood cake and releasing (extracting) proteins and nucleic acids which then can be processed for downstream analysis.

**Figure 1.**
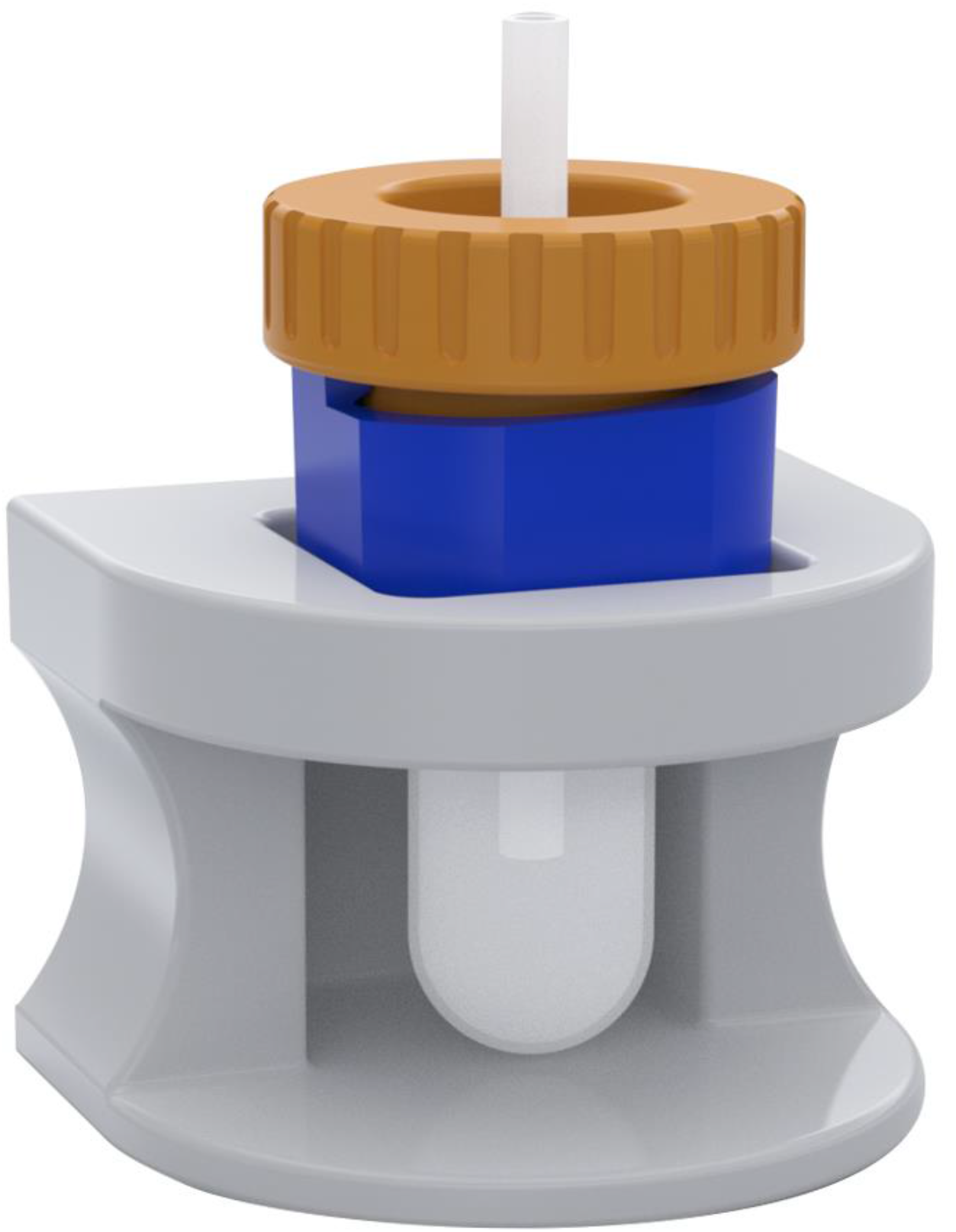
truCOLLECT Collection System. A digital drawing of the truCOLLECT collection tube and ergonomic tube holder. A protruding glass capillary allows for sample collection from a lanced finger into the tube containing an immobilization reagent.

In this study, antibodies against SARS-CoV-2 S- and N-protein were measured in 32 individuals. Blood samples were drawn by both venepuncture (followed by plasma preparation) and truCOLLECT Whole Blood (followed by on-demand plasma protein extraction via AFA), and titres were compared directly between methods.

## Materials and Methods

### Adaptive Focused Acoustics

Adaptive Focused Acoustics is the driving force behind sample extraction from blood collected with the truCOLLECT device. AFA is a highly adaptable method of ultrasonication whereby ultra-high frequency electronics and transducers produce and focus acoustic waves to effect change in a sample (Figure 2A). Energy fluctuations are manipulated to cause the formation of microscopic bubbles in dissolved gasses that grow, oscillate, and collapse (Figure 2B). This process results in localized pressure changes that can be adapted to gently mix samples, disrupt biomolecule complexes, or even fragment macro molecules. AFA treatment is completely non-contact which reduces the risk of contamination and offers a high degree of thermal control to maintain sample stability. Focused ultrasonicator instruments can be formatted to process specimens in single tube or plate formats, allowing for complete automation of workflows (Figure 2C).

**Figure 2.**
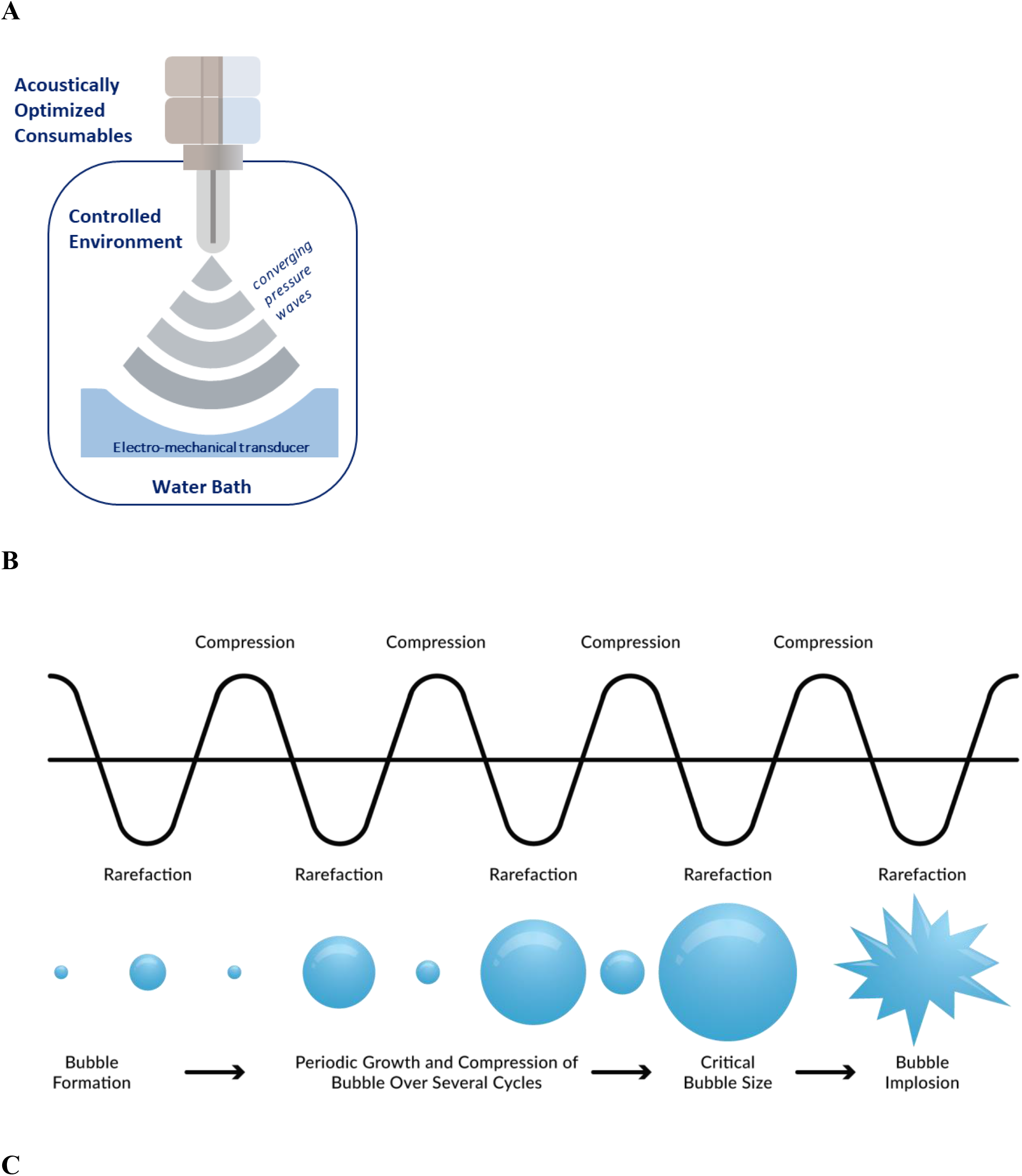
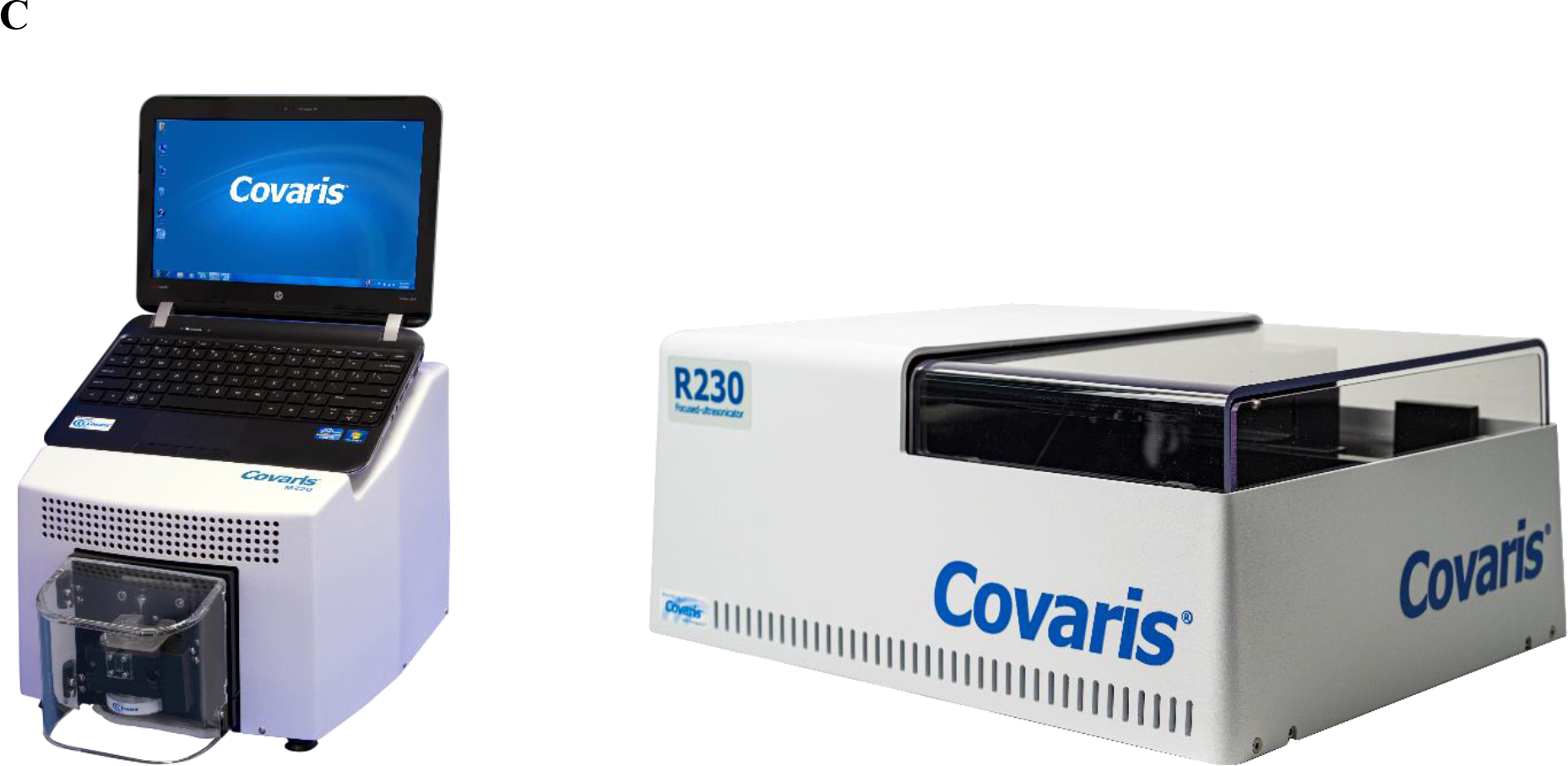
Adaptive Focused Acoustics Fundamentals. Transfer of acoustic energy from a focused-ultrasonicator transducer through a water bath and into a sample (A). Generation, oscillation, and collapse of microscopic bubbles in aqueous samples (B). The Covaris M220 (single tube; left) and R230 (plate format; right) focused-ultrasonicators (C).

### Blood Donor Demographics and Ethics

A cohort of 32 volunteers (Table 1) was recruited for this study, aged 20 to 67 years. The group consisted of 17 (53.1%) unvaccinated individuals and 15 (46.9%) people who received at least one dose of a SARS-CoV-2 mRNA vaccine. Positive tests previously verified that 17 (53.1%) participants had contracted and have since recovered from COVID-19. Of the unvaccinated volunteers, 9 (28.1%) have never tested positive for the virus based on personal communication. This study was approved by the Institutional Review Board (IRB) at Franciscan University of Steubenville (IRB #2022-05). All research was performed in accordance with relevant guidelines and regulations. Informed written consent was obtained from all participants.

**Table 1.**
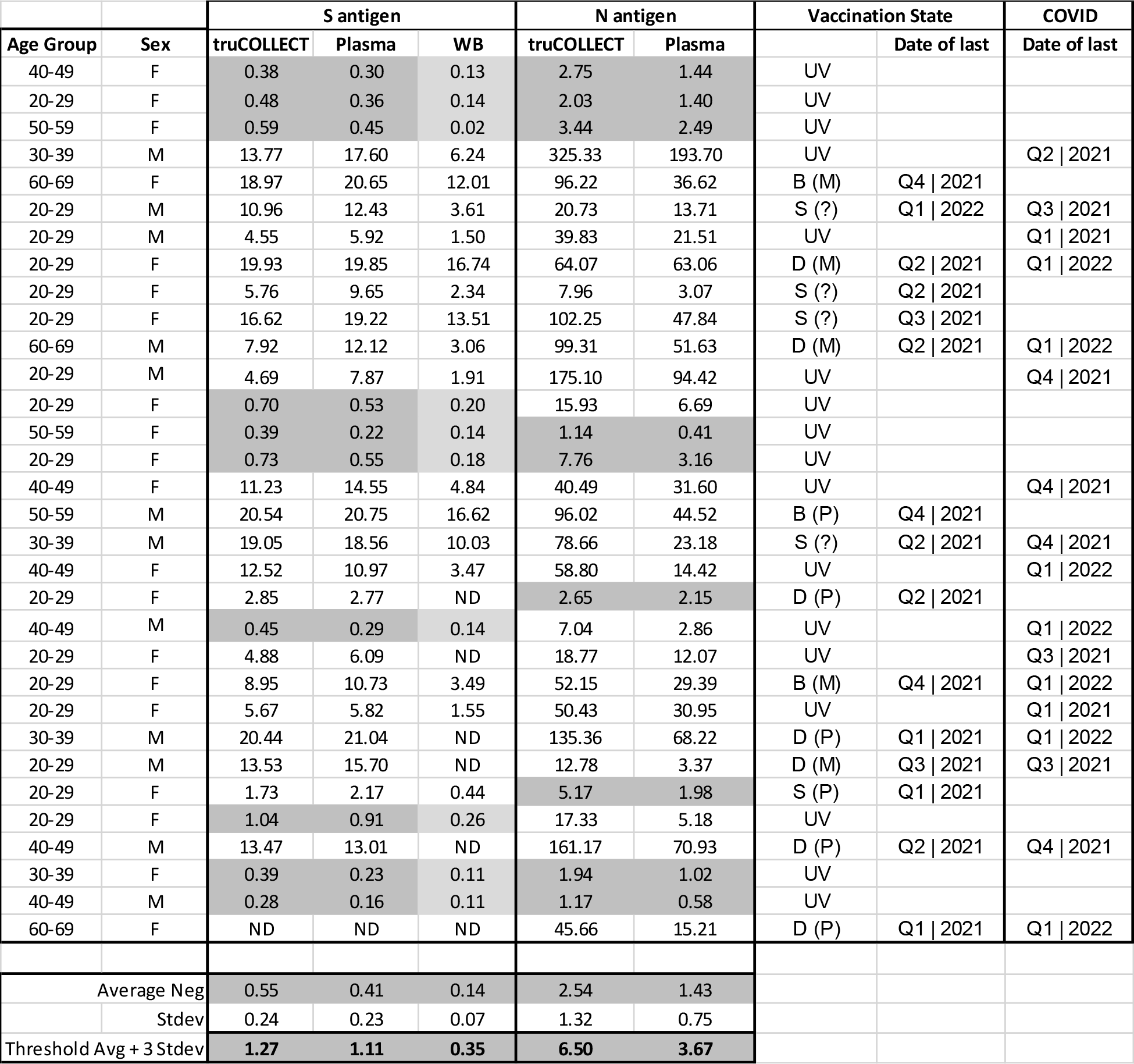
Donor Information. A table summarizing demographics, vaccination and SARS-CoV-2 infection status of blood donors, and ELISA data. Abbreviations: WB: Whole Blood (Fingerstick Microtainer); truCOLLECT: Whole Blood (Fingerstick truCOLLECT); Plasma: Plasma from Phlebotomy; M: Male; F: Female; S: single vaccination; D: double vaccination; B: Double vaccination + Booster; ã: unknown vaccination status; M/P/ã: Moderna/Pfizer/unspecified vaccine; UV: unvaccinated; ND: No Data; grey fields were used to calculate the NEG threshold (average negative value + 3 stdev).

| Age Group | Sex | S antigen |  |  | N antigen |  | Vaccination State |  | COVID |
| --- | --- | --- | --- | --- | --- | --- | --- | --- | --- |
|  |  | truCOLLECT | Plasma | WB | truCOLLECT | Plasma |  | Date of last | Date of last |
| 40-49 | F | 0.38 | 0.30 | 0.13 | 2.75 | 1.44 | UV |  |  |
| 20-29 | F | 0.48 | 0.36 | 0.14 | 2.03 | 1.40 | UV |  |  |
| 50-59 | F | 0.59 | 0.45 | 0.02 | 3.44 | 2.49 | UV |  |  |
| 30-39 | M | 13.77 | 17.60 | 6.24 | 325.33 | 193.70 | UV |  | Q2 \| 2021 |
| 60-69 | F | 18.97 | 20.65 | 12.01 | 96.22 | 36.62 | B (M) | Q4 \| 2021 |  |
| 20-29 | M | 10.96 | 12.43 | 3.61 | 20.73 | 13.71 | S (?) | Q1 \| 2022 | Q3 \| 2021 |
| 20-29 | M | 4.55 | 5.92 | 1.50 | 39.83 | 21.51 | UV |  | Q1 \| 2021 |
| 20-29 | F | 19.93 | 19.85 | 16.74 | 64.07 | 63.06 | D (M) | Q2 \| 2021 | Q1 \| 2022 |
| 20-29 | F | 5.76 | 9.65 | 2.34 | 7.96 | 3.07 | S (?) | Q2 \| 2021 |  |
| 20-29 | F | 16.62 | 19.22 | 13.51 | 102.25 | 47.84 | S (?) | Q3 \| 2021 |  |
| 60-69 | M | 7.92 | 12.12 | 3.06 | 99.31 | 51.63 | D (M) | Q2 \| 2021 | Q1 \| 2022 |
| 20-29 | M | 4.69 | 7.87 | 1.91 | 175.10 | 94.42 | UV |  | Q4 \| 2021 |
| 20-29 | F | 0.70 | 0.53 | 0.20 | 15.93 | 6.69 | UV |  |  |
| 50-59 | F | 0.39 | 0.22 | 0.14 | 1.14 | 0.41 | UV |  |  |
| 20-29 | F | 0.73 | 0.55 | 0.18 | 7.76 | 3.16 | UV |  |  |
| 40-49 | F | 11.23 | 14.55 | 4.84 | 40.49 | 31.60 | UV |  | Q4 \| 2021 |
| 50-59 | M | 20.54 | 20.75 | 16.62 | 96.02 | 44.52 | B (P) | Q4 \| 2021 |  |
| 30-39 | M | 19.05 | 18.56 | 10.03 | 78.66 | 23.18 | S (?) | Q2 \| 2021 | Q4 \| 2021 |
| 40-49 | F | 12.52 | 10.97 | 3.47 | 58.80 | 14.42 | UV |  | Q1 \| 2022 |
| 20-29 | F | 2.85 | 2.77 | ND | 2.65 | 2.15 | D (P) | Q2 \| 2021 |  |
| 40-49 | M | 0.45 | 0.29 | 0.14 | 7.04 | 2.86 | UV |  | Q1 \| 2022 |
| 20-29 | F | 4.88 | 6.09 | ND | 18.77 | 12.07 | UV |  | Q3 \| 2021 |
| 20-29 | F | 8.95 | 10.73 | 3.49 | 52.15 | 29.39 | B (M) | Q4 \| 2021 | Q1 \| 2022 |
| 20-29 | F | 5.67 | 5.82 | 1.55 | 50.43 | 30.95 | UV |  | Q1 \| 2021 |
| 30-39 | M | 20.44 | 21.04 | ND | 135.36 | 68.22 | D (P) | Q1 \| 2021 | Q1 \| 2022 |
| 20-29 | M | 13.53 | 15.70 | ND | 12.78 | 3.37 | D (M) | Q3 \| 2021 | Q3 \| 2021 |
| 20-29 | F | 1.73 | 2.17 | 0.44 | 5.17 | 1.98 | S (P) | Q1 \| 2021 |  |
| 20-29 | F | 1.04 | 0.91 | 0.26 | 17.33 | 5.18 | UV |  |  |
| 40-49 | M | 13.47 | 13.01 | ND | 161.17 | 70.93 | D (P) | Q2 \| 2021 | Q4 \| 2021 |
| 30-39 | F | 0.39 | 0.23 | 0.11 | 1.94 | 1.02 | UV |  |  |
| 40-49 | M | 0.28 | 0.16 | 0.11 | 1.17 | 0.58 | UV |  |  |
| 60-69 | F | ND | ND | ND | 45.66 | 15.21 | D (P) | Q1 \| 2021 | Q1 \| 2022 |
| Average Neg |  | 0.55 | 0.41 | 0.14 | 2.54 | 1.43 |  |  |  |
| Stdev |  | 0.24 | 0.23 | 0.07 | 1.32 | 0.75 |  |  |  |
| Threshold Avg + 3 Stdev |  | 1.27 | 1.11 | 0.35 | 6.50 | 3.67 |  |  |  |

### Venepuncture Blood Sample Collection and Processing

Venepuncture blood samples were collected by phlebotomists with K-Shield Advantage Winged Blood Collection Sets (Kawasumi) into EDTA Vacutainer Tubes (Becton Dickinson). A total volume of 6 mL was collected from each donor. Samples were processed by centrifugation of whole blood for 10 minutes at 2400 rpm and 4 C. Plasma was collected from the supernatant and stored at −80 C prior to analysis. Date of sample collection was between April 2022 and June 2022.

### Capillary Blood Sample Collection

The truCOLLECT Whole Blood Collection Kit (Covaris) was utilized to collect an exactly metered volume of capillary (fingerstick) blood via a 50 µL K_3_EDTA-coated capillary, which was then expelled onto an immobilization matrix in a closed vessel. The vessel also served as the extraction device, thereby minimizing sample loss and avoiding tracking issues. This kit contains an instruction manual (IFU), collection vessels, desiccant caps, gauze pads, alcohol wipes, disposable lancets, a pump dispenser, and adhesive bandages. Briefly, a contact-activated lancet was used to puncture the fingertip which was then applied directly to the capillary protruding from the collection device. After the capillary was filled completely, blood was dispensed into the collection tube and actively desiccated for 24 hours at ambient temperature. All truCOLLECT samples were stored until processing at ambient temperature.

As an additional comparator, capillary blood samples (from fingerstick) were also collected using Microtainer MAP Microtubes containing K_2_EDTA (Becton Dickinson). These whole blood samples were stored at room temperature prior to analysis. A workflow diagram for all sample types is presented in Figure 3.

**Figure 3.**
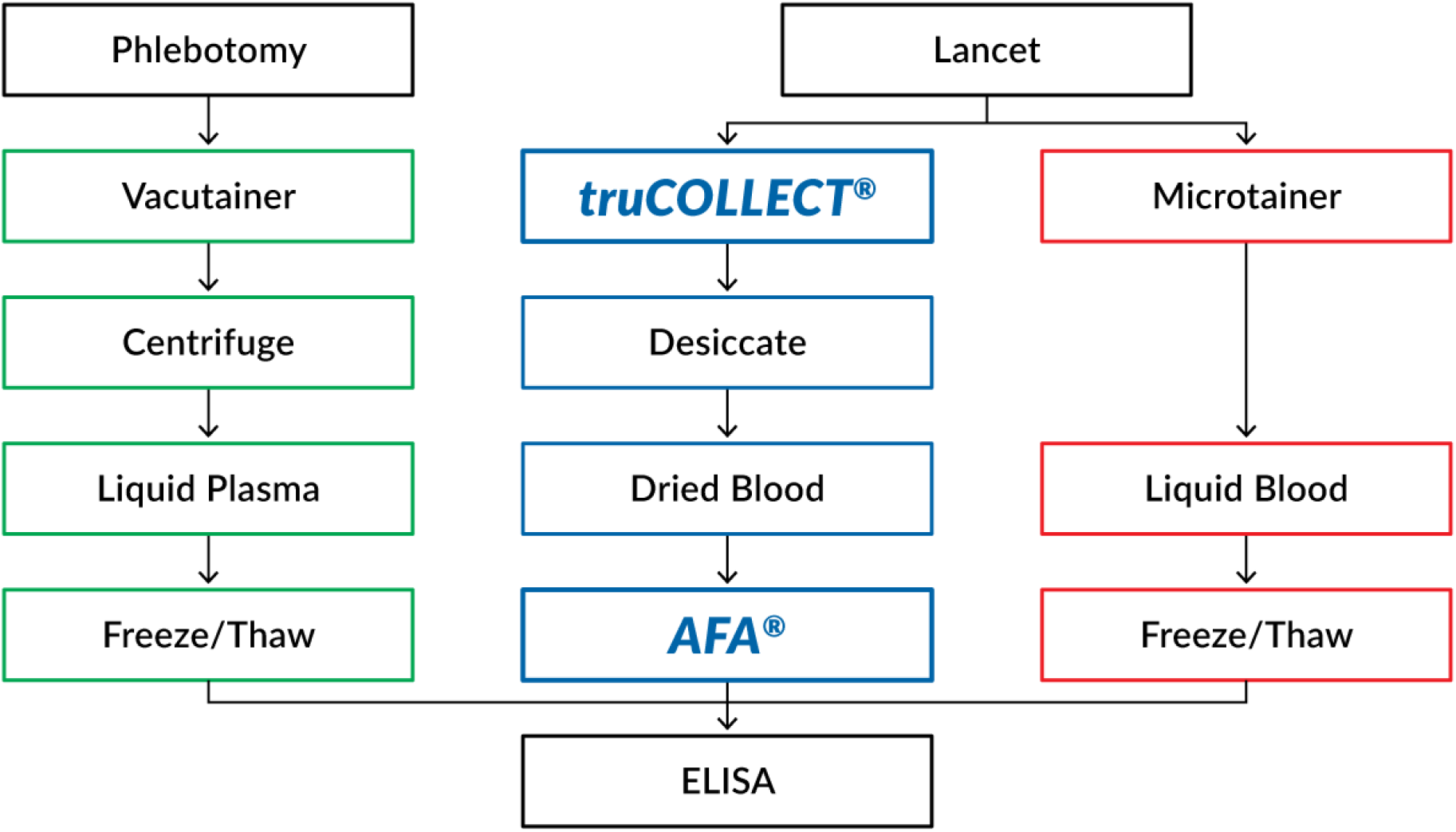
Experimental Workflow Diagram. A flowchart summarizing the three workflows tested for the processing of donor blood samples.

### truCOLLECT Sample Processing

Desiccated truCOLLECT whole blood samples were processed according to manufacturer protocol. After removal of the desiccant-containing cap, 350 µL of ELISA buffer (1X PBS, 0.05% Triton X-100) was added to the desiccated blood cake and subjected to AFA treatment in an M220 Focused-ultrasonicator (Covaris). The extraction process utilizes an average incident power of 52.5 J with a repetitive pulsing configuration. A method consisting of 40 pulsing iterations results in a total time of 5 minutes per sample. After AFA-treatment, samples were centrifuged for 5 minutes at 5,000 x g, and 300 µL supernatant was transferred to clean microcentrifuge tubes. All samples not used directly for ELISA were stored at −80 C prior to analysis. Frozen samples were thawed to room temperature and vortexed before running serological assays.

### Serology

Serological analysis was performed by ELISA assays targeting human IgG antibodies against SARS-CoV-2 S-protein and SARS-CoV-2 N-protein (Abcam, ab275300 and ab274339, respectively). The ELISA kits were designed for use with serum or plasma. All samples were diluted to the degree recommended by manufacturer protocols and included the appropriate controls and standards. The assays were run following manufacturer protocols, and absorbances were measured with a microplate reader (Tecan, Infinite M200pro). To compare results more accurately across data sets, threshold values were calculated from the average absorbances of confirmed negative donors plus 3 standard deviations.

### Analytical Analysis

Raw absorbance was generated by subtracting reference wavelength (570 nm) values from measurement wavelength (450 nm) values. Average absorbances were calculated from replicates for controls, standards, and samples. The COVID-19 S-Protein ELISA utilized a calibrator control to determine relative antibody concentrations, while the COVID-19 N-Protein ELISA employed a standard curve. Plasma data was plotted against both whole blood (fingerstick collected into Microtainer MAP Microtubes) and truCOLLECT Whole Blood data, and relationships were determined by calculating Pearson’s correlation coefficient with 95% confidence intervals. A direct comparison between absorbance values of different sample types was calculated by determining the ratio of paired donor samples.

## Results

### anti-SARS-CoV-2 Spike Protein Titre Analysis

ELISA analysis of donor samples resulted in the detection of antibodies against SARS-CoV-2 S-protein in 21 (67.7%) volunteers. Of the 10 donors testing negative, one had a self-identified case of COVID-19 in February 2022. Paired plasma data were plotted against truCOLLECT Whole Blood data (Figure 4), which resulted in a Pearson correlation coefficient of 0.98 CI [0.96, 0.98] (P-value <0.0001). The absorbance values of plasma and truCOLLECT were similar, with an average ratio of 1.06 (± 0.32). Paired plasma data were also plotted against whole blood (fingerstick Microtainer) data (Figure 5) to determine the correlation between sample types. This comparison resulted in a non-linear, but exponential correlation (Pearson correlation coefficient of 0.94 (P-value <0.0001)) indicating interference/inhibition at low titre levels in the whole blood samples as compared to plasma. In general, absorbances from whole blood samples were determined to be significantly lower than both plasma and truCOLLECT samples from paired donors.

**Figure 4.**
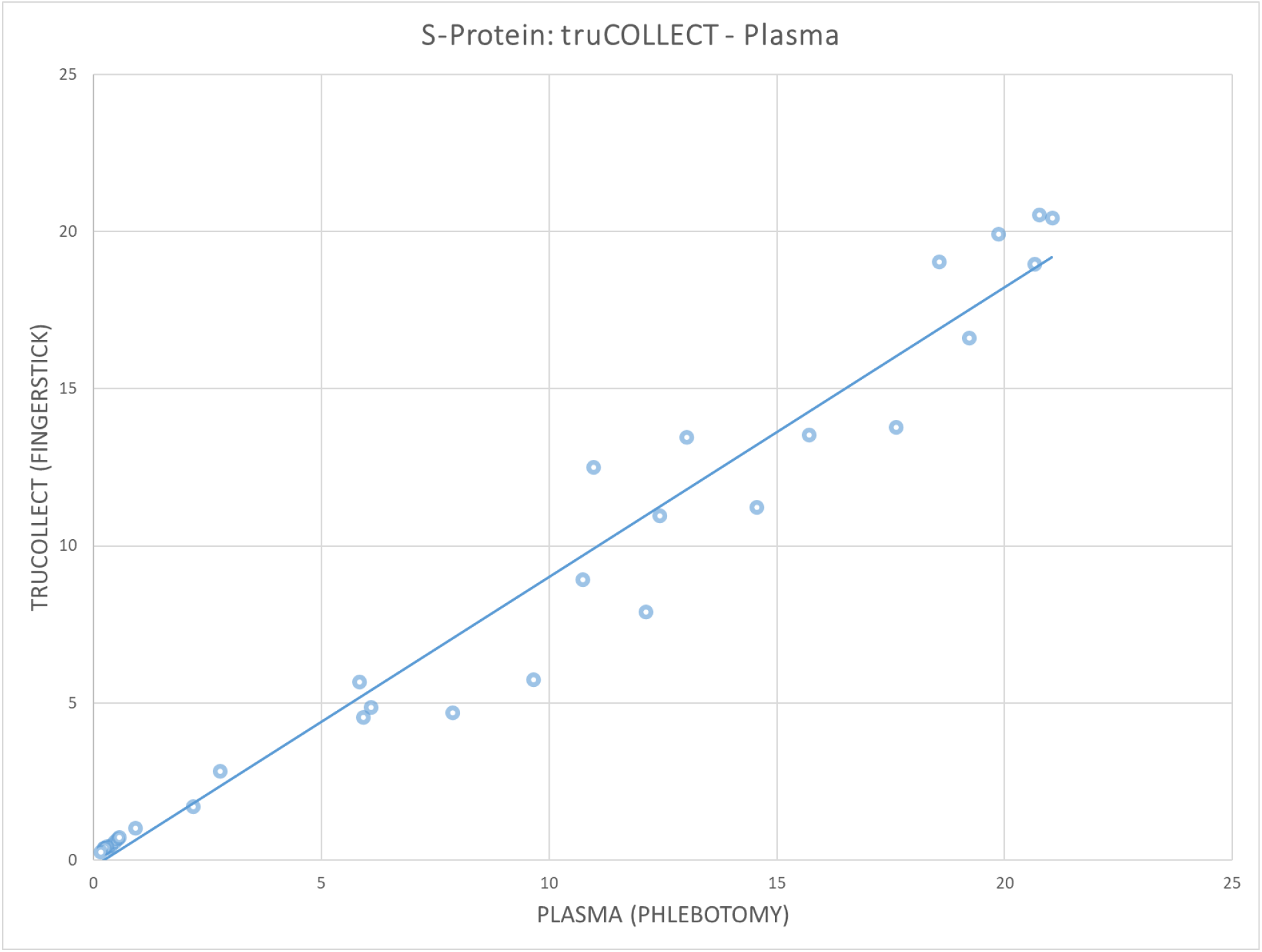
Correlation of SARS-CoV-2 S-protein antibodies in paired plasma and truCOLLECT samples. A comparison of absorbance measurements from 31 donors using the SARS-CoV-2 Spike Glycoprotein ELISA kit (Abcam). Capillary fingerstick blood collected with truCOLLECT (Y-axis), and Venepuncture derived plasma (X-axis) are examined. Trendline equation y = 0.9214X - 0.2153; r 0.981; 95% CI [0.96, 0.99]

**Figure 5.**
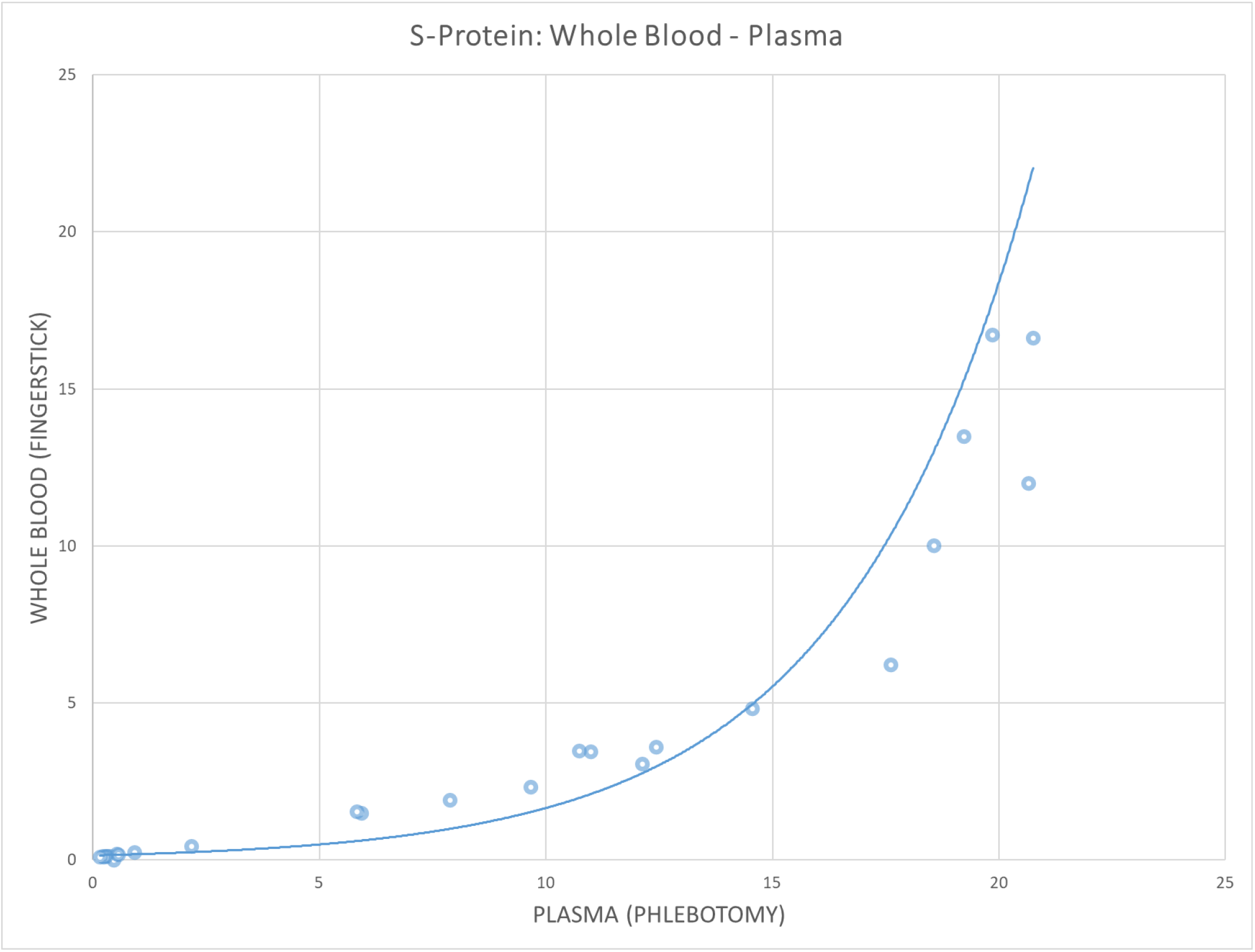
Correlation of SARS-CoV-2 S-protein antibodies in paired plasma and whole blood samples. A comparison of absorbance measurements from 31 donors using the SARS-CoV-2 Spike Glycoprotein ELISA kit (Abcam). Capillary fingerstick blood collected with Microtainer MAP Microtubes (Y-axis), and Venepuncture derived plasma (X-axis) are examined. Trendline equation y = 0.1498e^0.2405X^; r 0.898; 95% CI [0.78, 0.95]

### anti-SARS-CoV-2 Nucleocapsid Protein Titre Analysis

ELISA analysis of donor samples resulted in the detection of antibodies against SARS-CoV-2 N-protein in 23 (71.9%) volunteers using truCOLLECT Whole Blood lysate and 19 (59.4%) using plasma. Of the additional 4 positives detected in truCOLLECT-derived samples, one had a confirmed case of COVID-19 in September 2021, and another had a confirmed case in February 2022. A subset of 7 donors who tested positive in truCOLLECT-derived samples had previously reported no symptoms of COVID-19, nor tested positive. Paired plasma data were plotted against truCOLLECT data (Figure 6) to determine the correlation between sample types. The Pearson correlation coefficient was determined to be 0.97 (P value <0.001). Absorbance values for truCOLLECT were higher than plasma for all donors except 1 who tested negative for SARS-CoV-2 N-protein antibodies in both sample types.

**Figure 6.**
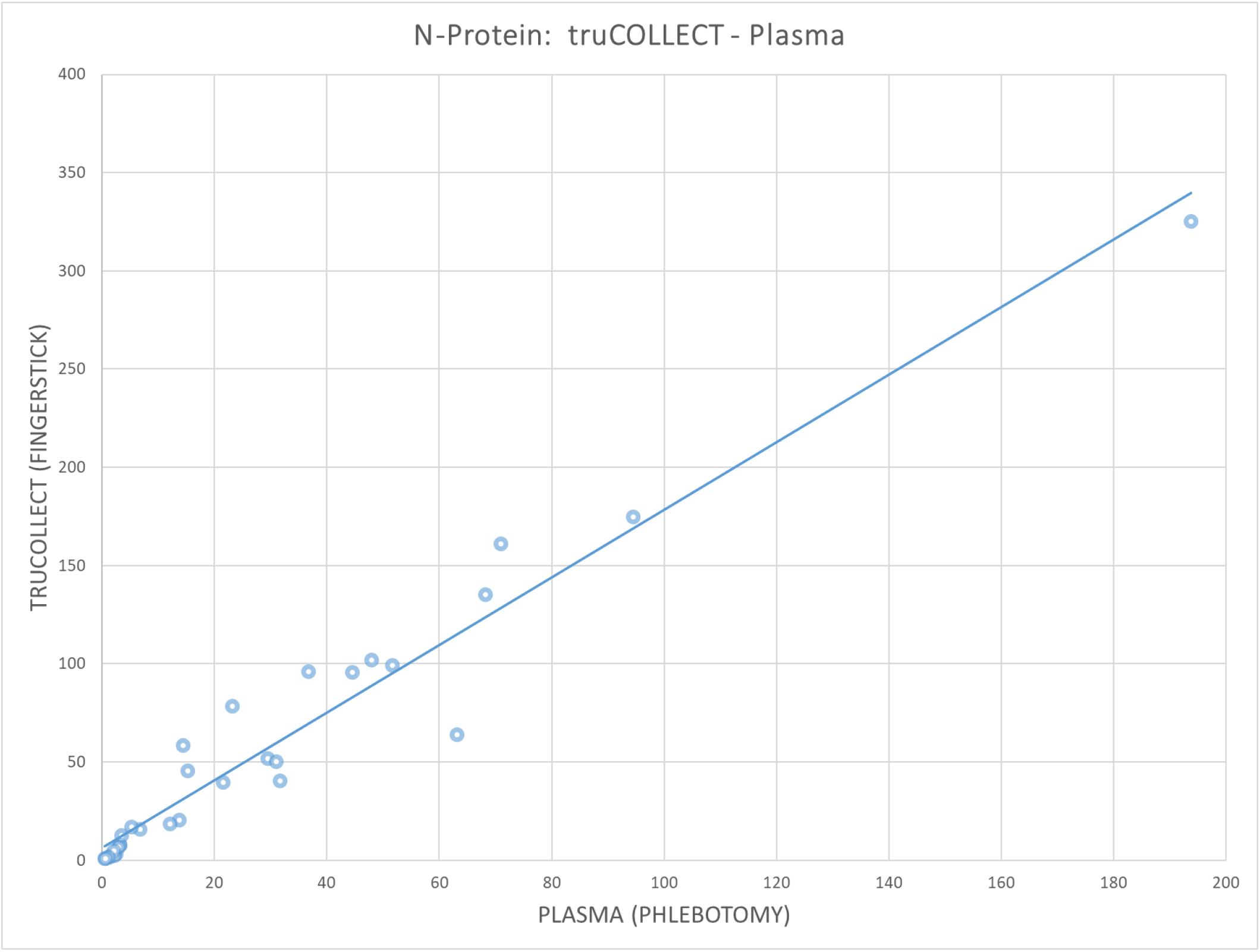
Correlation of SARS-CoV-2 N-protein antibodies in paired plasma and truCOLLECT samples. A comparison of absorbance measurements from 32 donors using the SARS-CoV-2 Nucleocapsid protein ELISA kit (Abcam). Capillary fingerstick blood collected with truCOLLECT (Y-axis), and Venepuncture derived plasma (X-axis) are examined. Trendline equation y = 1.7217X + 6.3161; r 0.972; 95% CI [0.94, 0.99]

## Discussion

The ELISA assays utilized herein were designed for use with serum or plasma. As such, the weak linear correlation between paired plasma and whole blood samples (Figure 5) can be expected due to the potential for interference with antibody binding as well as for high background absorbance. This is corroborated by the exponential fit suggesting interference/inhibition when using whole blood as ELISA input. Conversely, the strong linear correlation observed between paired plasma and truCOLLECT whole blood-derived samples (Figure 4) indicates that the lysate obtained from desiccated whole blood via AFA-enhanced rehydration and extraction using a non-denaturing buffer does not exhibit these inhibitory effects, and thereby closely mimics a plasma sample. A similarly high correlation between truCOLLECT lysates and plasma was observed for other plasma proteins, such as Dopamine β-hydroxylase (data not shown). Furthermore, anti-SARS-CoV-2 N-protein IgG titre measurements in truCOLLECT lysates indicated higher sensitivity in 31 (96.9%) volunteers as compared to paired plasma samples, suggesting efficient and quantitative AFA-based extraction of antibodies from desiccated whole blood. The increased sensitivity may be due to AFA-based active mixing of the resulting lysate leading to the dissociation of antibodies from non-specific targets/protein complexes, thereby improving availability for binding during the ELISA. The implications of these combined results are significant, suggesting a possible alternative to venepuncture for the collection of samples for antibody testing.

From truCOLLECT lysates we were able to accurately detect the humoral immunity against SARS-CoV-2 both for S and N proteins. When compared to self-reported patient immunity status to SARS-CoV-2 a subset of 7 (21.9%) vaccinated and unvaccinated individuals who declared no verifiable COVID-19 symptoms or exposures prior to this study tested positive for SARS-CoV-2 N-protein antibodies. This brings to light the growing need for rapid at-home collection of blood. Increased availability of simple self-collection options like truCOLLECT would significantly streamline serology testing which could enable more accurate disease monitoring strategies and help manage transmission throughout a population during a pandemic crisis.

The truCOLLECT kit offers a new avenue for serological testing. Accurate analysis of antibody titres currently requires travel to designated testing sites, long waiting periods, interaction with medical staff, and potential viral exposure. The truCOLLECT kit avoids these drawbacks while still allowing for highly accurate sample assessment by enabling decentralised collection of capillary blood that does not require cold chain transport and can be stored for extended periods prior to evaluation. This technology could serve to improve sample collection for a range of applications such as viral diagnostics, serological surveillance, and evaluating effectiveness of vaccines with large cohorts. In summary, our data suggests that capillary blood collection with truCOLLECT can serve as a viable alternative to conventional venepuncture for serological analysis.

## Data Availability

All data produced in the present study are available upon reasonable request to the authors.

## Acknowledgments

The authors would like to thank Eugenio Daviso (Covaris) for statistical analysis expertise.

